# Beyond the (log)book: Comparing accelerometer non-wear detection techniques in toddlers

**DOI:** 10.1101/2024.09.04.24312946

**Authors:** Elyse Letts, Sarah M da Silva, Natascja Di Cristofaro, Sara King-Dowling, Joyce Obeid

## Abstract

**Background:** Accelerometers are increasingly used to measure physical activity and sedentary time in toddlers. Data cleaning or wear time validation can impact outcomes of interest, particularly in young children who spend less time awake. However, no study has systematically compared wear time validation strategies in toddlers. As such, the objective of this study is to compare different fully-automated methods of distinguishing wear and non-wear time (counts and raw data algorithms) to the semi-automated (counts with logbooks) criterion method in toddlers.

**Methods:** We recruited 109 toddlers (age 12-35 mos) as part of the iPLAY study to wear an ActiGraph w-GT3X-BT accelerometer on the right hip for ∼7 consecutive days (removed for sleep and water activities). Parents completed a logbook to indicate monitor removal and nap times. We tested 15 nonwear detection methods grouped into 4 main categories: semi-automated logbook, consecutive 0 counts, modified consecutive 0 counts (Troiano and Choi), and raw data methods (van Hees and Ahmadi). Using logbooks as the criterion standard (all wear and wake-time only wear), we calculated the accuracy and F1 scores and compared overall wear time with a two one-sided test of equivalence.

**Results:** Participant daily wear time ranged from 556 to 684 minutes/day depending on method. Accuracy and F1 score ranged from 86 to 95%. Five methods were considered equivalent to the AllWear nonwear criterion (true wear time including sleep-time wear), with only one equivalent to the AwakeWear criterion. Mean absolute differences were lower for the AllWear criterion but ranged 49 to 192 minutes/day

**Conclusions:** The 5min0count, 10min_0count, 30min_0count, Troiano60s, and Ahmadi methods provide high accuracy and equivalency when compared to semi-automated cleaning using logbooks. This paper provides insights and quantitative results that can help researchers decide which method may be most appropriate given their population of interest, sample size, and study protocol.

## Introduction

Accelerometers have been increasingly used to measure physical activity and sedentary time in toddlers.^1–3^ A key element of analysis is wear time validation, which encompasses determining accelerometer wear and nonwear periods (e.g., removed for water activities, discomfort, etc.^4^) and differentiating wake-time vs. sleep-time wear.^5^ Existing evidence from school-age children and adults suggests that nonwear detection methods can significantly impact variables of interest,^5–7^ however, their impact in toddlers remains unclear.^3,8^

In school-age children, wear time may vary 17%, or up to 87 minutes per day, depending on method^6^ which could result in large overestimations of sedentary time.^4^ This is mirrored in adult studies, where nonwear methods can change daily wear time by up to 150 minutes.^9^ Further, in young children who nap regularly, determining nap wear time, so as not to overestimate sedentary time, is of greater concern.^10,11^ Given these large differences, it is important to clearly report the wear time validation methods used in toddlers.^5,6^

A recent review^8^ of accelerometry in the early years found that 1/3 of the 627 papers did not report nonwear detection methods, suggesting comprehensive/transparent reporting of methods remains a limitation. Of studies that did report methods, 20 minutes of consecutive 0 counts was most common.^8^ Other methods included logbooks (participant/proxy self-report), 1-120 minutes of consecutive 0 counts, and raw data variability.^8^

To date, a generally accepted criterion measure for determining nonwear time is logbooks. These logs allow the participant and/or caregiver proxy to note when the accelerometer was worn/removed each day. Despite being used as a criterion, they face inherent bias because they are self-reported. They also pose a significant participant burden and may not be completed accurately, leaving researchers to make a “best guess” based on observed count patterns. To address these limitations, logbooks are often used in conjunction with automated methods where data are manually checked against the logbooks to confirm the on/off times. Fully-automated methods of nonwear detection are growing, especially in large population-based studies as they can be used independently of logbooks, therefore reducing analysis burden.

One automated algorithm for nonwear detection is consecutive 0 counts. The basis of this method is that it is unlikely that an individual will remain completely motionless (0 counts) for a given amount of time (e.g., 60 minutes).^5^ Variations of this method change the total 0-count time (e.g., 20 minutes for younger children who are less likely to remain motionless) and/or allow for short-duration spikes of activity within the timeframe. Two further extensions of this method are integrated within ActiLife software (Troiano^12^ and Choi^13^), making them accessible and thus, commonly^7^ used. Finally, another growing category of automatic nonwear methods uses raw accelerations.^14^ While these methods may face challenges given the file size, data volume, and technical ability required to process the data, they have been consistently growing in use, but have limited testing in toddlers.

As outlined, there are a wide range of methods to determine nonwear time, all of which have been tested in adults^5,7^ and school-age children.^4,6^ However, to our knowledge, no study to date has compared nonwear detection methods in toddlers while accounting for potential nap-based wear,^15^ leaving researchers to use methods developed for older children or adapt adult methods. As such, this paper aims to answer the following question: What is the accuracy of existing automated accelerometer non-wear detection methods when compared to semi-automated logbook-based cleaning in toddlers?

## Methods

### Participants

We recruited healthy toddlers aged 12.0-35.9 months with no known physical disability, motor delay, or medical condition that affected movement, as part of the **i**nvestigating the validity and reliability of accelerometer-based measures of **P**hysica**L A**ctivity and sedentar**Y** time in toddlers (iPLAY) study. Participants were recruited through local childcare centres, websites/social media, and flyers throughout Hamilton, Ontario, Canada. A parent of each participant provided written informed consent. This study received ethics approval from the Hamilton Integrated Research Ethics Board (HiREB #3674).

### Study procedures

Participants were fitted with an ActiGraph wGT3X-BT (dynamic range ±8 *g*) by a member of the research team. The accelerometer was worn on the right hip (elastic belt) for approximately 7 consecutive days and removed only for night-time sleep and water activities. Parents were given the option to remove or keep the accelerometer on during daytime naps. All on/off times and nap times were recorded in a logbook (supplementary material 1 has a sample logbook). Accelerometers were initialized in ActiLife and recorded tri-axially at 30Hz with idle sleep mode activated. Upon accelerometer return, the raw .gt3x files, 1-second, and 60-second epoch AGD files were downloaded.

### Nonwear methods

We tested 15 different nonwear methods, broadly grouped into 4 categories: logbooks (criterion), consecutive 0 counts, modified consecutive 0 counts, and raw data algorithms. Table 1 provides further details on each method tested.

**Table 1.**
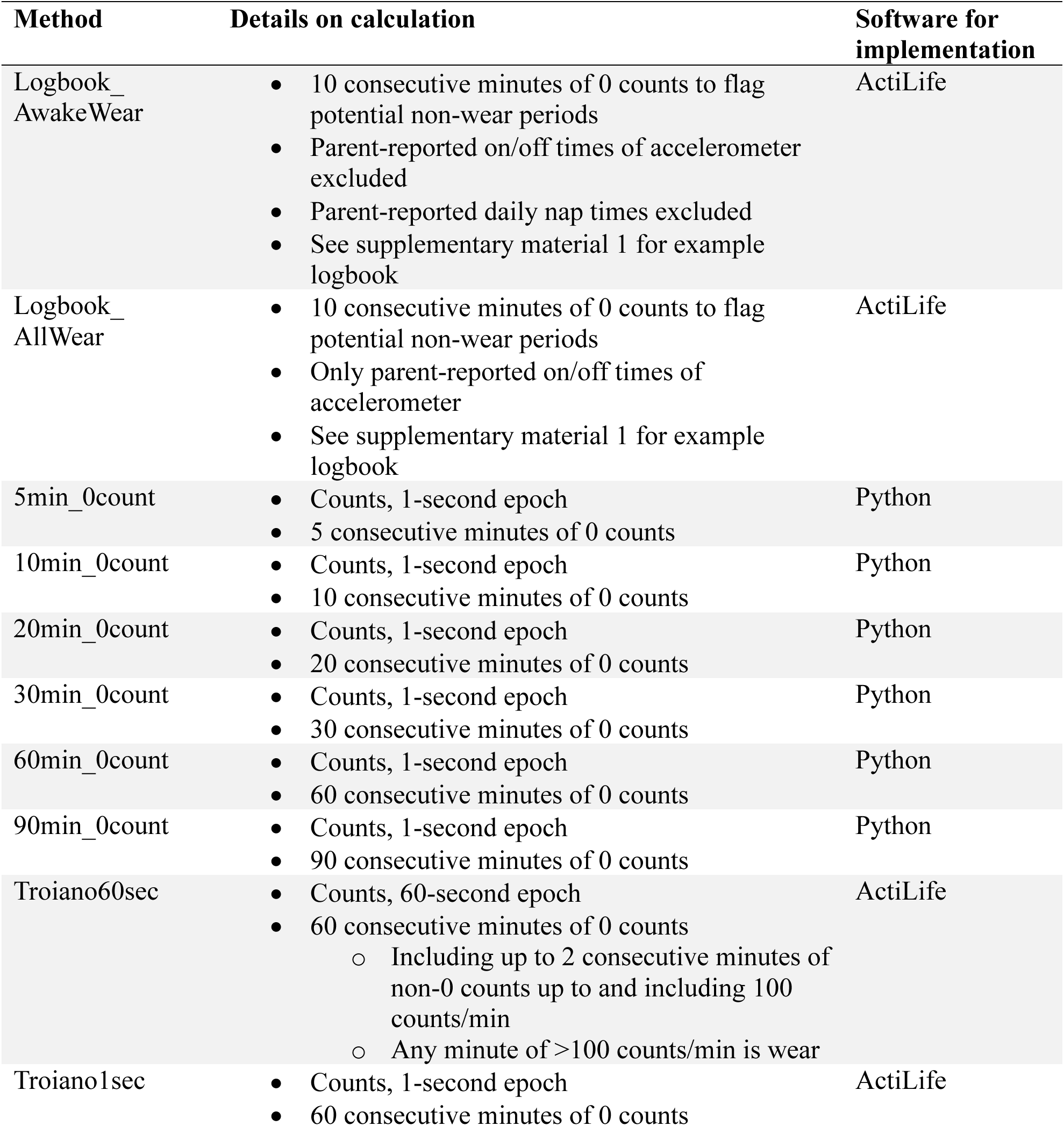
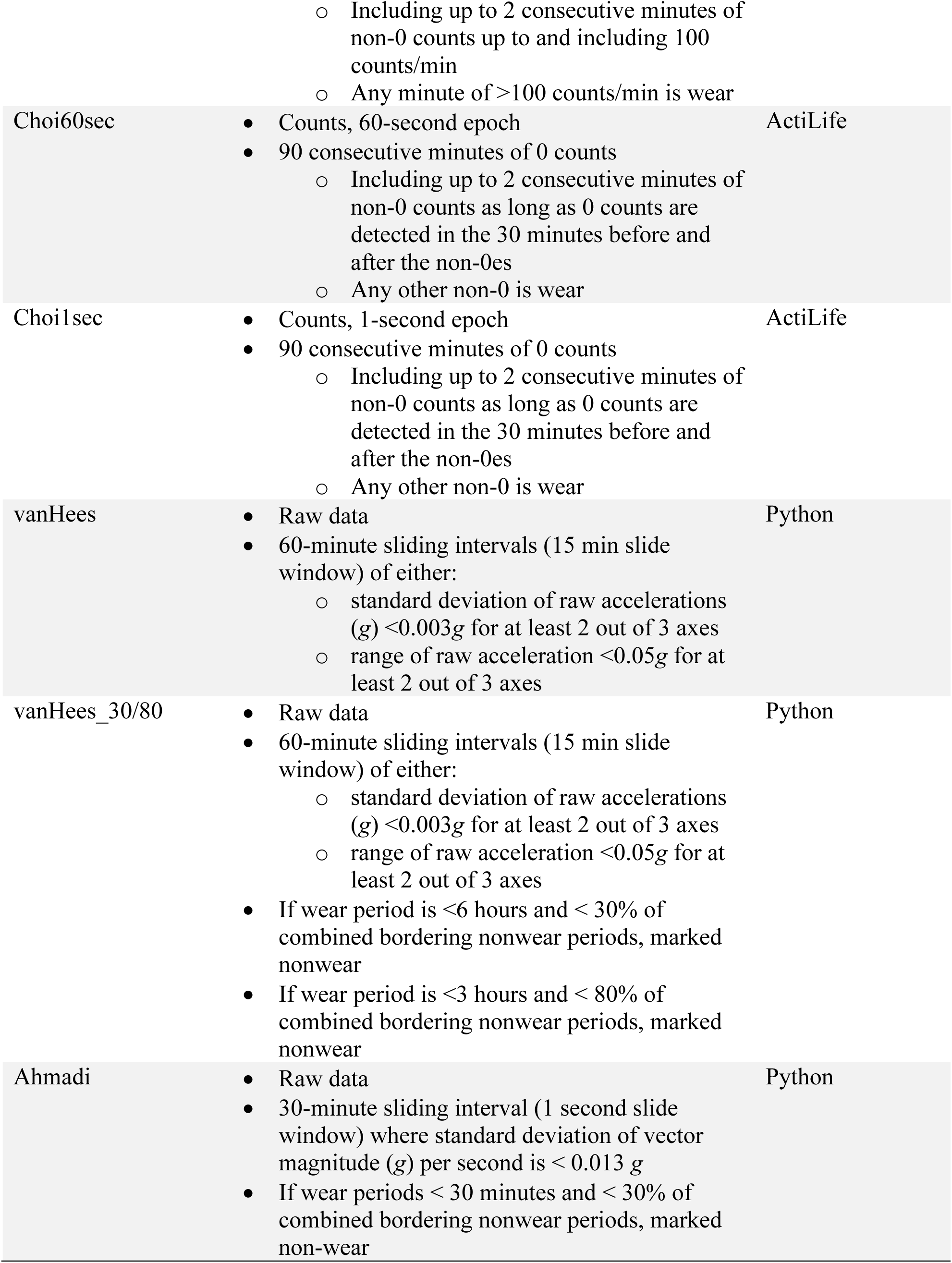
Details of nonwear detection methods compared in this study.

#### Logbook method (criterion)

The logbook method is a semi-automated protocol in which AGD files are compared to written logbooks. Using ActiLife software and 1-second AGD files, we identified potential nonwear using vector magnitude counts with a minimum nonwear length of 10 minutes, 0 spike tolerance, and ignored wear periods under 1 minute. The resulting wear/nonwear periods were then manually compared to logbooks. All nonwear periods noted in the logbook were removed using a 15-minute error window (e.g., participant notes device removal at 10:00 am, acceptable off-times could be from 9:45 to 10:15 am). If a nonwear period noted in the logbook was not detected by the software, a log diary was added to exclude it (e.g., participant removes the accelerometer for water activity, but a parent holds it, the software detects wear). If any on/off times were blank in the logbook, we used the time provided by ActiLife provided that it (1) was similar to other days in that week and (2) had a clear distinction between wear/nonwear. If the time provided by ActiLife did not meet these criteria, we used the first/last reported time (e.g., on time after nap) as the on/off time, respectively. Because parents were given the option to wear or remove the device during naps, we have elected to use two versions of logbook-based cleaning as criterion measures: 1) follows the logbook and leaves nap times in if accelerometer was worn during the nap (referred to as Logbook_AllWear hereafter); and 2) removes all nap times and only captures the time when the child is awake and wearing the accelerometer (referred to as Logbook_AwakeWear hereafter).

#### Consecutive 0 counts methods

The consecutive 0 count methods set a minimum required time of 0 counts to be considered nonwear. A count of 0 can be seen in both sedentary activity (e.g., no movement) and when the accelerometer is not worn (e.g., placed on a table). The theory is that it is unlikely for participants to be motionless for extended periods, suggesting the device is not being worn. Common minimum consecutive 0 counts range from 20 minutes (more common in children^8^), 60 minutes (more common in adults^17^), up to 120 minutes or more.^8,17^ Based on the findings from the 2022 review in the early years,^8^ we tested 5, 10, 20, 30, 60, and 90 minutes. All consecutive 0 count methods were implemented in Python, where .gt3x files were read with pygt3x(v0.5.2), and counts obtained using agcounts(v0.2.3).

#### Modified consecutive 0 counts methods

The modified consecutive 0 counts methods follow the same principle as the consecutive 0 count method, but add further “rules” around spike tolerances (non-0-count epochs surrounded by 0 counts), size of spikes, etc. Two common methods are available in ActiLife, and are developed by Troiano et al.^12^ (referred to as Troiano method hereafter) and Choi et al.^13^ (referred to as Choi method hereafter). The Troiano method was applied to the NHANES primary accelerometer analyses.^18^ Both methods were developed on a 60-second epoch count but are often applied to other epochs. We implemented both methods in ActiLife software using the count data in the original 60-second epoch and a 1-second epoch to allow for direct comparison to the other methods used in this paper.

#### Raw data methods

Raw data methods (using *g*’s, not counts) generally look at the variance of a signal within a given window and consider it nonwear if below a given threshold for that time window. These methods also often integrate additional “rules” related to short wear times surrounded by nonwear. The three methods tested here are those developed by van Hees et al.^19^ and by Ahmadi et al.^14^ (referred to as Ahmadi method hereafter). The van Hees method (used in GGIR^20^) uses a 3 m*g* standard deviation or 50 m*g* range threshold for 2 of the 3 axes using a 60-minute interval (sliding 15-minute window). It then checks the length of wear periods relative to bordering nonwear periods (see table 1 for details). We tested a previously published Python implementation of this method^7^ which computes only the thresholds part of the method (not the bordering nonwear periods; referred to as van Hees method hereafter). This method was validated against the original published R code. We also tested an adapted version of the code which incorporates the bordering nonwear period times criteria (see table 1 for details; referred to as van Hees 30/80 method hereafter). For both versions we read in the raw data from .gt3x files using pygt3x. The Ahmadi method uses a 13 m*g* vector magnitude threshold for a minimum of 30 minutes to determine nonwear, with bordering wear time rules (see table 1). We adapted the published R code to Python for use in this study and tested it against the R code.

### Statistical Analysis

For each method, we determined second-by-second wear/nonwear classification and day-level summary measures, the daily total minutes of wear/nonwear. We calculated the relative and absolute differences between fully automatic methods to both Logbook criteria. We also conducted a Two-One Sided Test (TOST) of equivalence with an equivalence bound of 5% of the average criterion minutes/day of wear. Further, we calculated epoch-level accuracy and an F1 score. Accuracy was calculated as the number of epochs correctly identified (compared to both criterion) divided by the total number of epochs. An F1 score is the harmonized mean of precision and recall. It is calculated as 2*True_Positives/(2*True_Positives+False_Positives+False_Negatives), giving a value between 0 and 1 where, like accuracy, higher is better.^21^ F1 score (weighted) was chosen as a metric in addition to accuracy as it can also account for the imbalance of labels (many cases of one label but very few cases of another). It is important to note that when there is a binary outcome (such as wear/nonwear), simple F1 score and accuracy are mathematically the same, while the weighted F1 can better account for the label imbalance. Accuracy and F1 score were calculated in Python(v3.9.6) using scikitlearn(v1.2.2) and TOST equivalence testing was completed in R(v4.3.1) using package TOSTER(v0.8.0).

## Results

Participant (n=109) and parent demographics are presented in Table 2. Participants had 6.9 average wear days (range: 3-9). All participants had completed logbooks, although 11 participants missed some on/off times (5 participants missing 1 on/off time, 4 missing 2 times, 1 missing 4 times, and 1 missing all off times). All participants had at least one nap, and 90 (83%) participants wore the accelerometer during at least one nap (426 total naps with accelerometer).

**Table 2.** Detailed participant characteristics.

| Participant characteristic | Overall n = 109 |
| --- | --- |
| Age (months) |  |
| Mean (SD) | 21.5 (7.0) |
| Range | 12.0-36.2 |
| Sex |  |
| Female | 56 (51%) |
| Male | 53 (49%) |
| Race/Ethnicity |  |
| White | 78 (72%) |
| Asian | 5 (5%) |
| Latin American | 3 (3%) |
| Arab | 1 (1%) |
| Multiracial | 22 (20%) |
| Home language |  |
| English | 98 (90%) |
| Other (Mandarin, Punjabi, Spanish, Portuguese, Arabic, Khmer, Italian) | 8 (8%) |
| Not reported | 3 (2%) |
| Marital status |  |
| Married | 86 (79%) |
| Living common-law or living with a partner | 16 (15%) |
| Single, never married | 4 (4%) |
| Separated | 1 (1%) |
| Not reported | 2 (2%) |
| Parent 1 highest education |  |
| College/University | 103 (94%) |
| High school | 4 (4%) |
| Not reported | 2 (2%) |
| Parent 2 highest education |  |
| College/University | 96 (88%) |
| High school | 6 (6%) |
| Not reported | 7 (6%) |

Wear time per day for methods are presented in Table 3 and ranged from 559 (Logbook_AwakeWear) to 684 min/day (van Hees). The wear time between Logbook_AwakeWear and Logbook_AllWear differed by 53.8 min/day. The results of the TOST equivalency tests are also presented in Table 3 (TOST details in supplementary material 2).

**Table 3.**
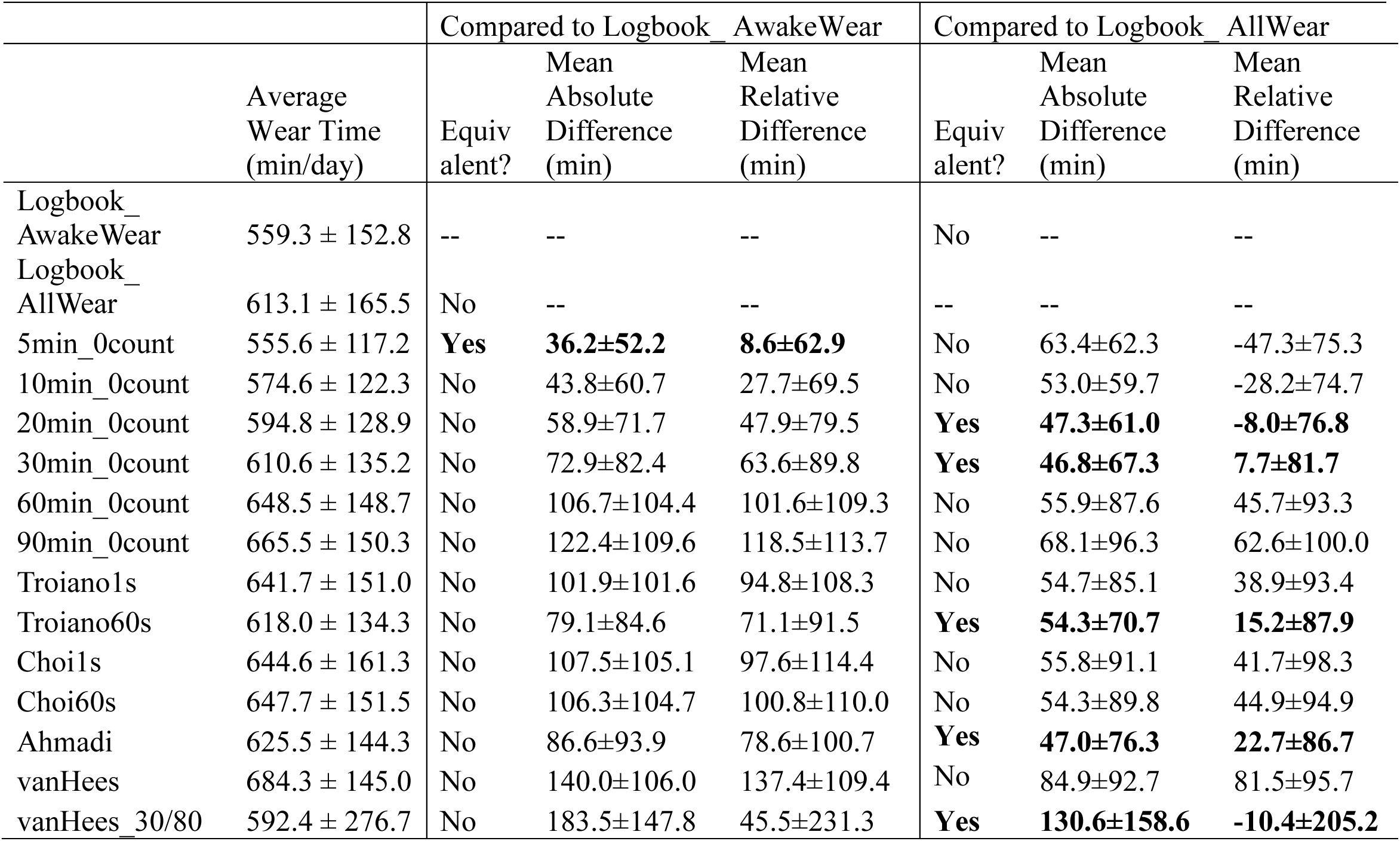
Wear time, TOST equivalency test results, and mean absolute and relative differences for each nonwear detection method. Values shown as mean±SD.

|  | Average<br>Wear Time<br>(min/day) | Compared to Logbook_AwakeWear |  |  | Compared to Logbook_AllWear |  |  |
| --- | --- | --- | --- | --- | --- | --- | --- |
|  |  | Equiv<br>alent? | Mean<br>Absolute<br>Difference<br>(min) | Mean<br>Relative<br>Difference<br>(min) | Equiv<br>alent? | Mean<br>Absolute<br>Difference<br>(min) | Mean<br>Relative<br>Difference<br>(min) |
| Logbook_AwakeWear | 559.3 ± 152.8 | -- | -- | -- | No | -- | -- |
| Logbook_AllWear | 613.1 ± 165.5 | No | -- | -- | -- | -- | -- |
| 5min_0count | 555.6 ± 117.2 | **Yes** | **36.2±52.2** | **8.6±62.9** | No | 63.4±62.3 | -47.3±75.3 |
| 10min_0count | 574.6 ± 122.3 | No | 43.8±60.7 | 27.7±69.5 | No | 53.0±59.7 | -28.2±74.7 |
| 20min_0count | 594.8 ± 128.9 | No | 58.9±71.7 | 47.9±79.5 | **Yes** | **47.3±61.0** | **-8.0±76.8** |
| 30min_0count | 610.6 ± 135.2 | No | 72.9±82.4 | 63.6±89.8 | **Yes** | **46.8±67.3** | **7.7±81.7** |
| 60min_0count | 648.5 ± 148.7 | No | 106.7±104.4 | 101.6±109.3 | No | 55.9±87.6 | 45.7±93.3 |
| 90min_0count | 665.5 ± 150.3 | No | 122.4±109.6 | 118.5±113.7 | No | 68.1±96.3 | 62.6±100.0 |
| Troiano1s | 641.7 ± 151.0 | No | 101.9±101.6 | 94.8±108.3 | No | 54.7±85.1 | 38.9±93.4 |
| Troiano60s | 618.0 ± 134.3 | No | 79.1±84.6 | 71.1±91.5 | **Yes** | **54.3±70.7** | **15.2±87.9** |
| Choi1s | 644.6 ± 161.3 | No | 107.5±105.1 | 97.6±114.4 | No | 55.8±91.1 | 41.7±98.3 |
| Choi60s | 647.7 ± 151.5 | No | 106.3±104.7 | 100.8±110.0 | No | 54.3±89.8 | 44.9±94.9 |
| Ahmadi | 625.5 ± 144.3 | No | 86.6±93.9 | 78.6±100.7 | **Yes** | **47.0±76.3** | **22.7±86.7** |
| vanHees | 684.3 ± 145.0 | No | 140.0±106.0 | 137.4±109.4 | No | 84.9±92.7 | 81.5±95.7 |
| vanHees_30/80 | 592.4 ± 276.7 | No | 183.5±147.8 | 45.5±231.3 | **Yes** | **130.6±158.6** | **-10.4±205.2** |

Methods considered equivalent to the Logbook_AllWear criterion were the 10min_0count, 30min_0count, Troiano60s, Ahmadi, and van Hees_30/80 methods. Compared to Logbook_AwakeWear, only 5min_0count was considered equivalent and had the smallest mean absolute difference in minutes of wear time. The mean absolute and relative differences per method are shown in Table 3 and Figure 1 (with distribution of relative differences). Compared to Logbook_AllWear, 30min_0count, Ahmadi, and 20min_0count methods had the smallest mean absolute differences.

**Figure 1.**
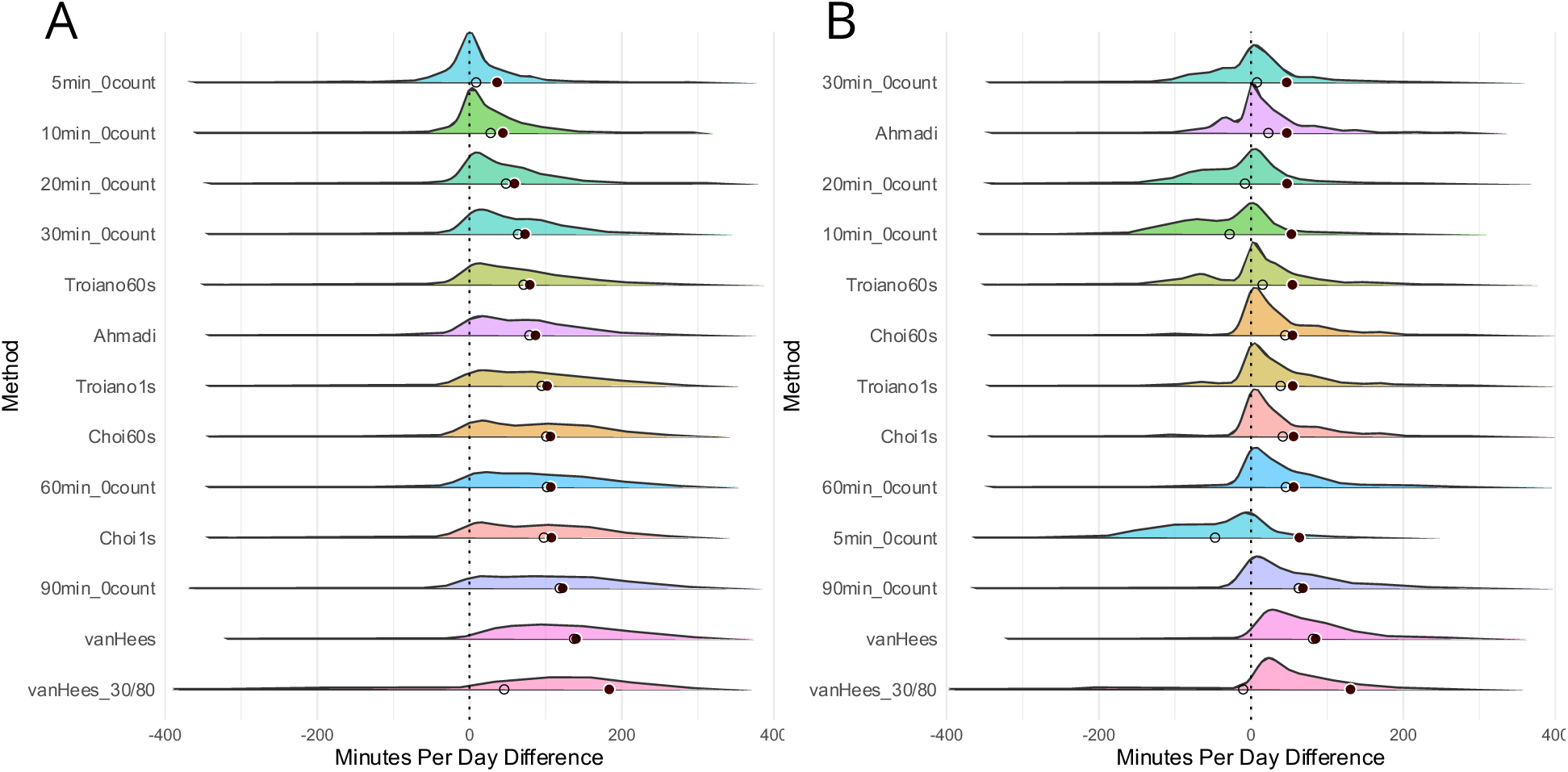
Absolute and relative differences in daily wear time for automatic methods compared to Logbook_AwakeWear (A) and Logbook_AllWear (B). Solid points represent the mean absolute differences, open circles represent the mean relative differences, and the half violin plots show the distribution of the by-day relative differences (can show whether a method tends to over-or under-estimate wear time).

The accuracy and F1 scores of methods compared to the logbooks are shown in Table 4. Compared to the Logbook_AllWear, Choi60s had the highest accuracy and F1 scores (95.1%) and vanHees_30/80 had the lowest (89.1%). Compared to Logbook_AwakeWear, 5min_0count (96.2%) and 10min_0count (96.1%) had the highest accuracy and F1 scores with van Hees_30/80 also having the lowest (86.1% accuracy, 86.2% F1 score). Figure 2 shows the average accuracy and accuracy per participant.

**Figure 2.**
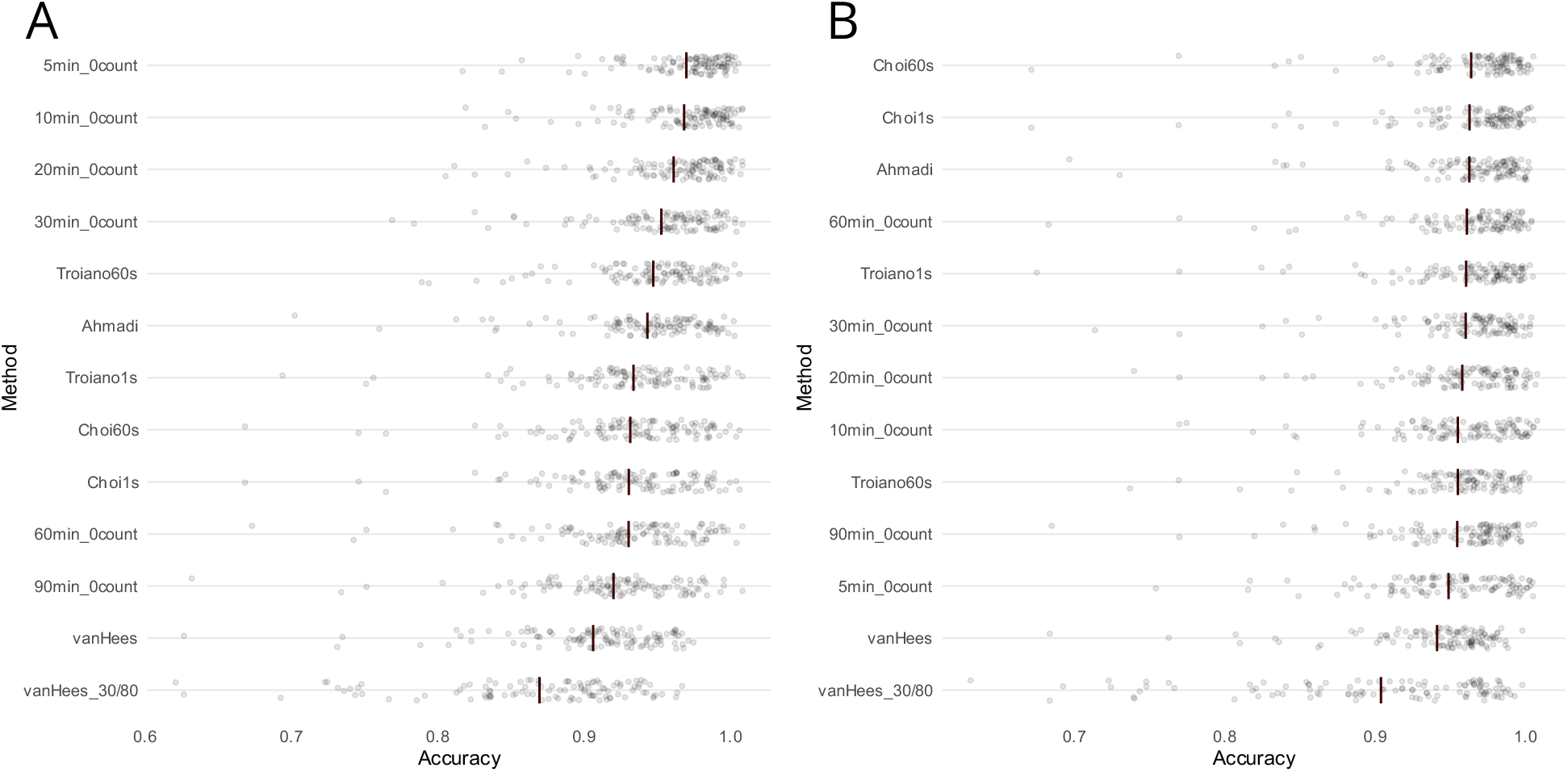
Accuracy for each method compared to Logbook_AwakeWear (A) and Logbook_AllWear (B). The solid line represents the mean accuracy, and the grey points are the accuracy for each participant.

**Table 4.** Accuracy and F1 scores of fully automatic methods compared to both Logbook criterion methods.

|  | Compared to Logbook_AwakeWear |  | Compared to Logbook_AllWear |  |
| --- | --- | --- | --- | --- |
|  | Accuracy | F1 Score | Accuracy | F1 Score |
| 5min_0count | 0.962 | 0.962 | 0.943 | 0.943 |
| 10min_0count | 0.961 | 0.961 | 0.949 | 0.949 |
| 20min_0count | 0.953 | 0.954 | 0.952 | 0.952 |
| 30min_0count | 0.945 | 0.945 | 0.955 | 0.955 |
| 60min_0count | 0.922 | 0.923 | 0.956 | 0.956 |
| 90min_0count | 0.911 | 0.912 | 0.949 | 0.949 |
| Troiano1s | 0.925 | 0.926 | 0.955 | 0.955 |
| Troiano60s | 0.939 | 0.940 | 0.949 | 0.949 |
| Choi1s | 0.922 | 0.923 | 0.957 | 0.958 |
| Choi60s | 0.923 | 0.924 | 0.958 | 0.959 |
| Ahmadi | 0.935 | 0.936 | 0.957 | 0.957 |
| vanHees | 0.898 | 0.899 | 0.936 | 0.936 |
| vanHees_30/80 | 0.861 | 0.862 | 0.898 | 0.898 |

## Discussion

Our findings demonstrate that the choice of nonwear detection method when using accelerometers in toddlers can alter wear-time outcomes, with methods differing by up to 129 min/day. Compared to Logbook_AwakeWear, most methods overestimated awake wear time and had slightly lower accuracy, while compared to Logbook_AllWear, methods more equally over– and under-estimated wear time and had higher accuracy. This is somewhat expected given that existing methods were designed to detect device removal, and excluding wear-time naps will decrease the total wear time – thus increasing the mean absolute difference and shifting the other methods to a relative overestimate. Overall, accuracy and F1 scores were relatively high (>85%) and varied very little for any given method, showing that the classes (wear/nonwear) were relatively balanced. When compared to Logbook_AwakeWear, only one method (5min_0count) was considered equivalent. Compared to Logbook_AllWear, more methods were equivalent, including mid-length consecutive zero counts methods and raw data methods with short wear periods. These novel results in toddlers generally support the findings from similar investigations in older children and adults, highlighting that method selection can have a meaningful impact on total wear time and provides valuable insights into wear-time considerations for toddlers.

In comparison to Logbook_AllWear, the Ahmadi raw data method was often highly ranked, having the lowest mean absolute difference and one of the highest accuracies (96%). Also highly accurate are the Choi methods, but interestingly they have larger mean absolute differences. For Logbook_AwakeWear, top-ranked were shorter 0count methods (5, 10, 20, and 30 minutes), with only the 5min_0count method considered equivalent. This suggests that sustained motionless time during napping is shorter than 10 mins. Both van Hees methods have the largest mean absolute differences and lowest accuracy and F1 scores when compared against both criterion methods. This aligns with previous findings in school-age children, which found that it had the largest mean bias and lowest agreement.^14^ Of note, the van Hees method was developed in adults and uses a relatively large window size (60 minutes) and low *g* threshold, which are not likely appropriate when applied to pediatric movement profiles.

Daily average awake wear is lower in toddlers than seen in older age groups,^6^ given their developmental increased sleep needs.^22^ The average awake-time wear reported in our study (559.3 min/day) is similar to other toddler studies with 547.2^23^ and 527.1^1^ min/day of wear time. As napping is almost ubiquitous in this age group, this highlights the importance of decision-making around naps. Logbook_AllWear looked at the “true” nonwear time, so included naps if the accelerometer was worn. However, as naps are difficult to detect when children are wearing the monitor,^10^ and there are currently no algorithms created to distinguish between naps and sedentary activities using hip-worn accelerometers,^15^ these nap times are likely to get misclassified as sedentary time. If sedentary time is the outcome of interest, researchers should consider asking parents to remove the device during naps, however, this could lead to a reduction in overall compliance as parents/caregivers may forget to put the device back on. Another solution could be our approach for Logbook_AwakeWear – have parents report whether the device was worn or not, and manually remove the worn nap times before further analysis. While time-consuming, this likely allows for the most accurate analysis. Alternatively, our results show that the 5min_0count method was equivalent to the Logbook_AwakeWear criterion, so could be considered when manual cleaning is not feasible. Ultimately, the chosen nonwear method may depend on the objective/context of a specific analysis. If the analysis is looking at total physical activity and sedentary time of toddlers with a small number of participants, the semi-automatic logbook method may be ideal, allowing for a more individualized nonwear detection. However, if it’s a larger study, or has mostly incomplete logbooks, a less subjective and less time/resource-intensive automated method may be preferred, including the 5min_0count, 30min_0count, and Ahmadi methods, depending on whether you are looking to match AwakeWear or AllWear.

Our study is not without limitations. First, as previously mentioned, logbooks were completed to different levels of accuracy/detail, so criterion measures may not exactly reflect the ground truth of accelerometer nonwear. Second, we also had a wide age range of toddlers (12-36 months), and the accuracy of accelerometer processing methods may vary based on individual-level characteristics (e.g. motor development, nap schedule). Finally, while we had a good representation of age and sex, participants were mostly White, English-speaking, dual-parent, highly educated families. As such, these results may not be generalizable to more diverse and lower-income families, who may experience more challenges with accelerometer wear and logbook completion.^24^ Logbooks provide a rich source of data when completed fully but add to participant burden which may be limiting for participants. An important avenue for future work should be to improve nap time vs. sedentary time detection in toddlers, as this is currently limited,^10,15^ and understanding which techniques work best across the wide developmental spectrum of toddlerhood.

## Conclusion

This study presents a novel analysis of nonwear detection methods for toddler accelerometry. The toddler age poses specific issues relating to nonwear detection and data cleaning (e.g., accounting for naps, reduced wake-time). While many methods were considered equivalent to the AllWear nonwear criterion, fewer were equivalent to AwakeWear. This paper provides important insights and quantitative results that can help researchers decide which accelerometry processing method may be best given their population of interest, sample size, and study protocol.

## Data availability statement

Data may be available upon request to the corresponding author.

## Ethics approval statement

This study received ethics clearance from the Hamilton integrated Research Ethics Board (HiREB #3674).

## Patient consent statement

Informed written consent was obtained from a parent of each participant.

## Acknowledgements

We would like to thank the participants and their families for their time and effort, as well as the funders who made this research possible. We also acknowledge the efforts of Ms. Camilla Wegrzynowska who contributed to the manual cleaning of accelerometer data.

