## Supplementary material for "Beyond the (log)book: Comparing accelerometer non-wear detection techniques in toddlers"

**Supplementary Material 1 – Sample logbook**

*Note.* This is a sample logbook written by the research team which contains no participant data.

**ACTIVITY DIARY:**

In addition to having your child wear the activity monitor for one week, we ask that you keep this diary to monitor the times the activity monitor was put on or taken off. This will help us to understand your child's regular physical activity. Please bring this form along with your child's activity monitor to your next study visit.

* at grandparents *

| Event | Example<br>Tues<br>June 19 | DAY 1<br>Day: Mon<br>Date: May 16 | DAY 2<br>Day: Tues<br>Date: May 17 | DAY 3<br>Day: Wed<br>Date: May 18 | DAY 4<br>Day: Thurs<br>Date: May 19 | DAY 5<br>Day: Fri<br>Date: May 20 | DAY 6<br>Day: Sat<br>Date: May 21 | DAY 7<br>Day: Sun<br>Date: May 22 |
|---|---|---|---|---|---|---|---|---|
| Time the device was put on | 7:10 am | ~7:00 am | 7:15 am | 7:45am | 6:15am | 6:30 am | 6:50 am | 7:10 am |
| Was the device removed for naps? | ☑ Yes<br>☐ No | ☑ Yes<br>☐ No | ☐ Yes<br>☑ No | ☐ Yes<br>☑ No | ☑ Yes<br>☐ No | ☐ Yes<br>☑ No | ☑ Yes<br>☐ No | ☑ Yes<br>☐ No |
| Nap Time (Please include both start of nap & wake up time) | 2:30 pm – 3:47 pm | 12:30 – 2:45 pm | 1:00 – 3:00 pm | | 12:45 – 2:30 pm | 1:15 – 3:30pm | 12:30 – 2:30 pm | 1:00 – 3:00 pm<br>* car sleep? |
| | - | | | | * forgot to put device back on | | | |
| Times the device may have been taken off and put back on and reason(s) (e.g. bath, swimming, etc) | 4:45 pm – 5:27 pm (swimming) | ~6:30 pm bath | 6:00 – 6:15 pm bath | 11:00 – 3:00pm baby bjorn * carried | 5:45 – 6:15pm bath ↓ | 11:00 – 11:30am car ride | 8:00 – 8:15 am change | 7:30 – 7:45am change |
| | 7:10 pm – 7:37 pm (bath) | | | | | 6:00 – 6:30pm bath | 4:00 – 4:15 pm swim | 1:00 – 3:00pm car ride |
| | - | 3:00 – 3:20 pm car ride | | 4:30 – 5:00 pm swim | | | ~6:00 pm bath | 6:00 – 6:25pm bath |
| | - | | | ~5:00 pm bath | | | | |
| Time the device was taken off before bed | 7:40 pm | 7:00 pm | 8:00pm | 6:30 pm | 6:30 pm | 6:45pm | 6:45pm | 7:00 pm |

ID #: Participant 1 Initials: P1 Accelerometer #: XXX / 01

**PLEASE SEE THE NEXT PAGE FOR DETAILED INSTRUCTIONS**

**Supplementary material 2. Results of TOST equivalency tests.**

**Compared to Logbook_AllWear**
[1] "==========="
[1] "LogbookAwakeWear_W"

Paired t-test

The equivalence test was non-significant, t(752) = -9.7, p = 1
The null hypothesis test was significant, t(752) = -21.5p < 0.01
NHST: reject null significance hypothesis that the effect is equal to zero
TOST: don't reject null equivalence hypothesis

TOST Results

| | t | df | p.value |
|---|---|---|---|
| t-test | -21.484 | 752 | < 0.001 |
| TOST Lower | -9.703 | 752 | 1 |
| TOST Upper | -33.264 | 752 | < 0.001 |

Effect Sizes

| | Estimate | SE | C.I. | Conf. Level |
|---|---|---|---|---|
| Raw | -55.9050 | 2.60218 | [-60.1905, -51.6195] | 0.9 |
| Hedges's g(z) | -0.7821 | 0.04164 | [-0.8504, -0.7135] | 0.9 |

Note: SMD confidence intervals are an approximation. See vignette("SMD_calcs").
[1] "==========="
[1] "Ahmadi_W"

Paired t-test

The equivalence test was significant, t(752) = -2.5, p < 0.01
The null hypothesis test was significant, t(752) = 7.194p < 0.01
NHST: reject null significance hypothesis that the effect is equal to zero
TOST: reject null equivalence hypothesis

TOST Results

| | t | df | p.value |
|---|---|---|---|
| t-test | 7.194 | 752 | < 0.001 |
| TOST Lower | 16.897 | 752 | < 0.001 |
| TOST Upper | -2.509 | 752 | 0.006 |

Effect Sizes

| | Estimate | SE | C.I. | Conf. Level |
|---|---|---|---|---|
| Raw | 22.7292 | 3.15947 | [17.5259, 27.9324] | 0.9 |
| Hedges's g(z) | 0.2619 | 0.03706 | [0.2009, 0.3227] | 0.9 |

Note: SMD confidence intervals are an approximation. See vignette("SMD_calcs").
[1] "==========="

[1] "vanHees_W"

Paired t-test

The equivalence test was non-significant, t(752) = 14.56, p = 1
The null hypothesis test was significant, t(752) = 23.351p < 0.01
NHST: reject null significance hypothesis that the effect is equal to zero
TOST: don't reject null equivalence hypothesis

TOST Results

```
            t  df p.value
t-test     23.35 752 < 0.001
TOST Lower 32.14 752 < 0.001
TOST Upper 14.56 752       1
```

Effect Sizes

```
             Estimate      SE              C.I. Conf. Level
Raw           81.4597 3.48842 [75.7146, 87.2047]        0.9
Hedges's g(z)  0.8501 0.04252     [0.78, 0.9198]        0.9
```

Note: SMD confidence intervals are an approximation. See vignette("SMD_calcs").
[1] "=========="
[1] "5min_0count_W"

Paired t-test

The equivalence test was non-significant, t(752) = -6, p = 1
The null hypothesis test was significant, t(752) = -17.2p < 0.01
NHST: reject null significance hypothesis that the effect is equal to zero
TOST: don't reject null equivalence hypothesis

TOST Results

```
              t  df p.value
t-test     -17.213 752 < 0.001
TOST Lower  -6.048 752       1
TOST Upper -28.378 752 < 0.001
```

Effect Sizes

```
             Estimate      SE                C.I. Conf. Level
Raw          -47.2602 2.74559 [-51.7819, -42.7386]        0.9
Hedges's g(z) -0.6267 0.03986     [-0.692, -0.561]        0.9
```

Note: SMD confidence intervals are an approximation. See vignette("SMD_calcs").
[1] "=========="
[1] "10min_0count _W"

Paired t-test

The equivalence test was non-significant, t(752) = 0.91, p = 0.18
The null hypothesis test was significant, t(752) = -10.4p < 0.01
NHST: reject null significance hypothesis that the effect is equal to zero
TOST: don't reject null equivalence hypothesis

TOST Results
t df p.value
t-test -10.3509 752 < 0.001
TOST Lower 0.9131 752 0.181
TOST Upper -21.6149 752 < 0.001

Effect Sizes
Estimate SE C.I. Conf. Level
Raw -28.1699 2.72150 [-32.6519, -23.6879] 0.9
Hedges's g(z) -0.3768 0.03771 [-0.4387, -0.3147] 0.9
Note: SMD confidence intervals are an approximation. See vignette("SMD_calcs").
[1] "=========="
[1] "20min_0count _W"

Paired t-test

The equivalence test was significant, t(752) = 8.08, p < 0.01
The null hypothesis test was significant, t(752) = -2.87p < 0.01
NHST: reject null significance hypothesis that the effect is equal to zero
TOST: reject null equivalence hypothesis

TOST Results
t df p.value
t-test -2.869 752 0.004
TOST Lower 8.082 752 < 0.001
TOST Upper -13.819 752 < 0.001

Effect Sizes
Estimate SE C.I. Conf. Level
Raw -8.0310 2.79941 [-12.6413, -3.4207] 0.9
Hedges's g(z) -0.1044 0.03654 [-0.1645, -0.0444] 0.9
Note: SMD confidence intervals are an approximation. See vignette("SMD_calcs").
[1] "=========="
[1] "30min_0count_W"

Paired t-test

The equivalence test was significant, t(752) = -7.7, p < 0.01
The null hypothesis test was significant, t(752) = 2.602p < 0.01
NHST: reject null significance hypothesis that the effect is equal to zero
TOST: reject null equivalence hypothesis

```
TOST Results
                 t  df p.value
t-test      2.602 752   0.009
TOST Lower 12.904 752 < 0.001
TOST Upper -7.700 752 < 0.001

Effect Sizes
              Estimate      SE              C.I. Conf. Level
Raw            7.74203 2.97566 [2.8415, 12.6426]         0.9
Hedges's g(z)  0.09472 0.03652  [0.0347, 0.1547]         0.9
Note: SMD confidence intervals are an approximation. See vignette("SMD_calcs").
[1] "==========="
[1] "60min_0count_W"

Paired t-test

The equivalence test was non-significant, t(752) = 4.43, p = 1
The null hypothesis test was significant, t(752) = 13.443p < 0.01
NHST: reject null significance hypothesis that the effect is equal to zero
TOST: don't reject null equivalence hypothesis

TOST Results
                 t  df p.value
t-test     13.443 752 < 0.001
TOST Lower 22.459 752 < 0.001
TOST Upper  4.427 752       1

Effect Sizes
              Estimate      SE              C.I. Conf. Level
Raw            45.7063 3.40007 [40.1068, 51.3058]         0.9
Hedges's g(z)   0.4894 0.03856   [0.4259, 0.5526]         0.9
Note: SMD confidence intervals are an approximation. See vignette("SMD_calcs").
[1] "==========="
[1] "90min_0count_W"

Paired t-test

The equivalence test was non-significant, t(752) = 8.78, p = 1
The null hypothesis test was significant, t(752) = 17.188p < 0.01
NHST: reject null significance hypothesis that the effect is equal to zero
TOST: don't reject null equivalence hypothesis

TOST Results
                 t  df p.value
t-test     17.188 752 < 0.001
```

```
TOST Lower 25.599 752 < 0.001
TOST Upper  8.777 752       1

Effect Sizes
            Estimate      SE              C.I. Conf. Level
Raw           62.6426 3.64461 [56.6403, 68.6448]         0.9
Hedges's g(z)   0.6257 0.03985      [0.56, 0.691]        0.9
Note: SMD confidence intervals are an approximation. See vignette("SMD_calcs").
[1] "=========="
[1] "vanHees_30/80_W"

Paired t-test

The equivalence test was significant, t(752) = 2.71, p < 0.01
The null hypothesis test was non-significant, t(752) = -1.39p = 0.17
NHST: don't reject null significance hypothesis that the effect is equal to zero
TOST: reject null equivalence hypothesis

TOST Results
              t  df p.value
t-test     -1.387 752   0.166
TOST Lower  2.712 752   0.003
TOST Upper -5.486 752 < 0.001

Effect Sizes
             Estimate      SE              C.I. Conf. Level
Raw          -10.37302 7.47906 [-22.6902, 1.9441]         0.9
Hedges's g(z)  -0.05049 0.03647  [-0.1104, 0.0094]         0.9
Note: SMD confidence intervals are an approximation. See vignette("SMD_calcs").
[1] "=========="
[1] "Choi1s_W"

Paired t-test

The equivalence test was non-significant, t(752) = 3.1, p = 1
The null hypothesis test was significant, t(752) = 11.653p < 0.01
NHST: reject null significance hypothesis that the effect is equal to zero
TOST: don't reject null equivalence hypothesis

TOST Results
              t  df p.value
t-test     11.653 752 < 0.001
TOST Lower 20.210 752 < 0.001
TOST Upper  3.095 752   0.999

Effect Sizes
```

| | Estimate | SE | C.I. | Conf. Level |
|---|---|---|---|---|
| Raw | 41.7422 | 3.58223 | [35.8427, 47.6417] | 0.9 |
| Hedges's g(z) | 0.4242 | 0.03805 | [0.3616, 0.4866] | 0.9 |

Note: SMD confidence intervals are an approximation. See vignette("SMD_calcs").
[1] "==========="
[1] "Choi60s_W"

Paired t-test

The equivalence test was non-significant, t(752) = 4.11, p = 1
The null hypothesis test was significant, t(752) = 12.981p < 0.01
NHST: reject null significance hypothesis that the effect is equal to zero
TOST: don't reject null equivalence hypothesis

TOST Results

| | t | df | p.value |
|---|---|---|---|
| t-test | 12.981 | 752 | < 0.001 |
| TOST Lower | 21.849 | 752 | < 0.001 |
| TOST Upper | 4.114 | 752 | 1 |

Effect Sizes

| | Estimate | SE | C.I. | Conf. Level |
|---|---|---|---|---|
| Raw | 44.8767 | 3.45703 | [39.1834, 50.5701] | 0.9 |
| Hedges's g(z) | 0.4726 | 0.03842 | [0.4093, 0.5356] | 0.9 |

Note: SMD confidence intervals are an approximation. See vignette("SMD_calcs").
[1] "==========="
[1] "Troiano1s_W"

Paired t-test

The equivalence test was non-significant, t(752) = 2.41, p = 0.99
The null hypothesis test was significant, t(752) = 11.413p < 0.01
NHST: reject null significance hypothesis that the effect is equal to zero
TOST: don't reject null equivalence hypothesis

TOST Results

| | t | df | p.value |
|---|---|---|---|
| t-test | 11.413 | 752 | < 0.001 |
| TOST Lower | 20.415 | 752 | < 0.001 |
| TOST Upper | 2.412 | 752 | 0.992 |

Effect Sizes

| | Estimate | SE | C.I. | Conf. Level |
|---|---|---|---|---|
| Raw | 38.8680 | 3.40551 | [33.2596, 44.4765] | 0.9 |
| Hedges's g(z) | 0.4155 | 0.03798 | [0.353, 0.4778] | 0.9 |

Note: SMD confidence intervals are an approximation. See vignette("SMD_calcs").

[1] "==========="
[1] "Troiano60s_W"

Paired t-test

The equivalence test was significant, t(752) = -4.8, p < 0.01
The null hypothesis test was significant, t(752) = 4.736p < 0.01
NHST: reject null significance hypothesis that the effect is equal to zero
TOST: reject null equivalence hypothesis

TOST Results

| | t | df | p.value |
|---|---|---|---|
| t-test | 4.736 | 752 | < 0.001 |
| TOST Lower | 14.306 | 752 | < 0.001 |
| TOST Upper | -4.834 | 752 | < 0.001 |

Effect Sizes

| | Estimate | SE | C.I. | Conf. Level |
|---|---|---|---|---|
| Raw | 15.1695 | 3.20321 | [9.8942, 20.4449] | 0.9 |
| Hedges's g(z) | 0.1724 | 0.03671 | [0.112, 0.2327] | 0.9 |

Note: SMD confidence intervals are an approximation. See vignette("SMD_calcs").

**Compared to Logbook_AwakeWear**
[1] "==========="
[1] "LogbookAllWear_W"

Paired t-test

The equivalence test was non-significant, t(752) = 10.74, p = 1
The null hypothesis test was significant, t(752) = 21.484p < 0.01
NHST: reject null significance hypothesis that the effect is equal to zero
TOST: don't reject null equivalence hypothesis

TOST Results

| | t | df | p.value |
|---|---|---|---|
| t-test | 21.48 | 752 | < 0.001 |
| TOST Lower | 32.23 | 752 | < 0.001 |
| TOST Upper | 10.74 | 752 | 1 |

Effect Sizes

| | Estimate | SE | C.I. | Conf. Level |
|---|---|---|---|---|
| Raw | 55.9050 | 2.60218 | [51.6195, 60.1905] | 0.9 |
| Hedges's g(z) | 0.7821 | 0.04164 | [0.7135, 0.8504] | 0.9 |

Note: SMD confidence intervals are an approximation. See vignette("SMD_calcs").
[1] "==========="
[1] "Ahmadi_W"

Paired t-test

The equivalence test was non-significant, t(752) = 13.81, p = 1
The null hypothesis test was significant, t(752) = 21.431p < 0.01
NHST: reject null significance hypothesis that the effect is equal to zero
TOST: don't reject null equivalence hypothesis

TOST Results

| | t | df | p.value |
|---|---|---|---|
| t-test | 21.43 | 752 | < 0.001 |
| TOST Lower | 29.05 | 752 | < 0.001 |
| TOST Upper | 13.81 | 752 | 1 |

Effect Sizes

| | Estimate | SE | C.I. | Conf. Level |
|---|---|---|---|---|
| Raw | 78.6342 | 3.66921 | [72.5914, 84.6769] | 0.9 |
| Hedges's g(z) | 0.7802 | 0.04162 | [0.7116, 0.8484] | 0.9 |

Note: SMD confidence intervals are an approximation. See vignette("SMD_calcs").
[1] "==========="
[1] "vanHees_W"

Paired t-test

The equivalence test was non-significant, t(752) = 27.45, p = 1
The null hypothesis test was significant, t(752) = 34.464p < 0.01
NHST: reject null significance hypothesis that the effect is equal to zero
TOST: don't reject null equivalence hypothesis

TOST Results

| | t | df | p.value |
|---|---|---|---|
| t-test | 34.46 | 752 | < 0.001 |
| TOST Lower | 41.48 | 752 | < 0.001 |
| TOST Upper | 27.45 | 752 | 1 |

Effect Sizes

| | Estimate | SE | C.I. | Conf. Level |
|---|---|---|---|---|
| Raw | 137.365 | 3.98579 | [130.8005, 143.9288] | 0.9 |
| Hedges's g(z) | 1.255 | 0.04872 | [1.1743, 1.3345] | 0.9 |

Note: SMD confidence intervals are an approximation. See vignette("SMD_calcs").

```
[1] "=========="
[1] "5min_0count_W"

Paired t-test

The equivalence test was significant, t(752) = -8.4, p < 0.01
The null hypothesis test was significant, t(752) = 3.77p < 0.01
NHST: reject null significance hypothesis that the effect is equal to zero
TOST: reject null equivalence hypothesis

TOST Results
              t  df p.value
t-test      3.770 752 < 0.001
TOST Lower 15.965 752 < 0.001
TOST Upper -8.425 752 < 0.001

Effect Sizes
           Estimate      SE              C.I. Conf. Level
Raw           8.6448 2.29311 [4.8683, 12.4213]        0.9
Hedges's g(z)   0.1372 0.03661   [0.077, 0.1974]        0.9
Note: SMD confidence intervals are an approximation. See vignette("SMD_calcs").
[1] "=========="
[1] "10min_0count_W"

Paired t-test

The equivalence test was non-significant, t(752) = -0.091, p = 0.46
The null hypothesis test was significant, t(752) = 10.944p < 0.01
NHST: reject null significance hypothesis that the effect is equal to zero
TOST: don't reject null equivalence hypothesis

TOST Results
                t  df p.value
t-test     10.94411 752 < 0.001
TOST Lower 21.97892 752 < 0.001
TOST Upper -0.09071 752   0.464

Effect Sizes
           Estimate      SE               C.I. Conf. Level
Raw          27.7351 2.53425 [23.5615, 31.9087]        0.9
Hedges's g(z)   0.3984 0.03786   [0.3361, 0.4605]        0.9
Note: SMD confidence intervals are an approximation. See vignette("SMD_calcs").
[1] "=========="
[1] "20min_0count_W"

Paired t-test
```

The equivalence test was non-significant, t(752) = 6.87, p = 1
The null hypothesis test was significant, t(752) = 16.524p < 0.01
NHST: reject null significance hypothesis that the effect is equal to zero
TOST: don't reject null equivalence hypothesis

TOST Results

| | t | df | p.value |
|---|---|---|---|
| t-test | 16.524 | 752 | < 0.001 |
| TOST Lower | 26.176 | 752 | < 0.001 |
| TOST Upper | 6.872 | 752 | 1 |

Effect Sizes

| | Estimate | SE | C.I. | Conf. Level |
|---|---|---|---|---|
| Raw | 47.8740 | 2.8973 | [43.1025, 52.6455] | 0.9 |
| Hedges's g(z) | 0.6016 | 0.0396 | [0.5363, 0.6665] | 0.9 |

Note: SMD confidence intervals are an approximation. See vignette("SMD_calcs").
[1] "=========="
[1] "30min_0count_W"

Paired t-test

The equivalence test was non-significant, t(752) = 10.91, p = 1
The null hypothesis test was significant, t(752) = 19.459p < 0.01
NHST: reject null significance hypothesis that the effect is equal to zero
TOST: don't reject null equivalence hypothesis

TOST Results

| | t | df | p.value |
|---|---|---|---|
| t-test | 19.46 | 752 | < 0.001 |
| TOST Lower | 28.01 | 752 | < 0.001 |
| TOST Upper | 10.91 | 752 | 1 |

Effect Sizes

| | Estimate | SE | C.I. | Conf. Level |
|---|---|---|---|---|
| Raw | 63.6471 | 3.27090 | [58.2603, 69.0339] | 0.9 |
| Hedges's g(z) | 0.7084 | 0.04076 | [0.6412, 0.7752] | 0.9 |

Note: SMD confidence intervals are an approximation. See vignette("SMD_calcs").
[1] "=========="
[1] "60min_0count_W"

Paired t-test

The equivalence test was non-significant, t(752) = 18.49, p = 1
The null hypothesis test was significant, t(752) = 25.504p < 0.01
NHST: reject null significance hypothesis that the effect is equal to zero

```
TOST: don't reject null equivalence hypothesis

TOST Results
             t  df p.value
t-test     25.50 752 < 0.001
TOST Lower 32.52 752 < 0.001
TOST Upper 18.49 752       1

Effect Sizes
            Estimate      SE               C.I. Conf. Level
Raw           101.6113 3.98410 [95.0499, 108.1726]         0.9
Hedges's g(z)   0.9285 0.04359    [0.8566, 0.9999]         0.9
Note: SMD confidence intervals are an approximation. See vignette("SMD_calcs").
[1] "=========="
[1] "90min_0count_W"

Paired t-test

The equivalence test was non-significant, t(752) = 21.86, p = 1
The null hypothesis test was significant, t(752) = 28.613p < 0.01
NHST: reject null significance hypothesis that the effect is equal to zero
TOST: don't reject null equivalence hypothesis

TOST Results
             t  df p.value
t-test     28.61 752 < 0.001
TOST Lower 35.36 752 < 0.001
TOST Upper 21.86 752       1

Effect Sizes
            Estimate      SE                C.I. Conf. Level
Raw            118.548 4.14310 [111.7244, 125.3708]         0.9
Hedges's g(z)    1.042 0.04526      [0.967, 1.1158]         0.9
Note: SMD confidence intervals are an approximation. See vignette("SMD_calcs").
[1] "=========="
[1] "vanHees_30/80_W"

Paired t-test

The equivalence test was non-significant, t(752) = 2.08, p = 0.98
The null hypothesis test was significant, t(752) = 5.402p < 0.01
NHST: reject null significance hypothesis that the effect is equal to zero
TOST: don't reject null equivalence hypothesis

TOST Results
             t  df p.value
```

```
t-test     5.402 752 < 0.001
TOST Lower 8.719 752 < 0.001
TOST Upper 2.084 752   0.981

Effect Sizes
           Estimate      SE             C.I. Conf. Level
Raw          45.5320 8.42914 [31.6502, 59.4138]        0.9
Hedges's g(z)   0.1967 0.03679    [0.1361, 0.257]        0.9
Note: SMD confidence intervals are an approximation. See vignette("SMD_calcs").
[1] "==========="
[1] "Choi1s_W"

Paired t-test

The equivalence test was non-significant, t(752) = 16.71, p = 1
The null hypothesis test was significant, t(752) = 23.423p < 0.01
NHST: reject null significance hypothesis that the effect is equal to zero
TOST: don't reject null equivalence hypothesis

TOST Results
            t  df p.value
t-test     23.42 752 < 0.001
TOST Lower 30.13 752 < 0.001
TOST Upper 16.71 752       1

Effect Sizes
           Estimate      SE             C.I. Conf. Level
Raw          97.6472 4.16894 [90.7815, 104.513]        0.9
Hedges's g(z)   0.8527 0.04255   [0.7825, 0.9224]        0.9
Note: SMD confidence intervals are an approximation. See vignette("SMD_calcs").
[1] "==========="
[1] "Choi60s_W"

Paired t-test

The equivalence test was non-significant, t(752) = 18.16, p = 1
The null hypothesis test was significant, t(752) = 25.138p < 0.01
NHST: reject null significance hypothesis that the effect is equal to zero
TOST: don't reject null equivalence hypothesis

TOST Results
            t  df p.value
t-test     25.14 752 < 0.001
TOST Lower 32.11 752 < 0.001
TOST Upper 18.16 752       1
```

Effect Sizes

| | Estimate | SE | C.I. | Conf. Level |
|---|---|---|---|---|
| Raw | 100.7818 | 4.00914 | [94.1792, 107.3843] | 0.9 |
| Hedges's g(z) | 0.9152 | 0.04341 | [0.8436, 0.9863] | 0.9 |

Note: SMD confidence intervals are an approximation. See vignette("SMD_calcs").
[1] "==========="
[1] "Troiano1s_W"

Paired t-test

The equivalence test was non-significant, t(752) = 16.93, p = 1
The null hypothesis test was significant, t(752) = 24.02p < 0.01
NHST: reject null significance hypothesis that the effect is equal to zero
TOST: don't reject null equivalence hypothesis

TOST Results

| | t | df | p.value |
|---|---|---|---|
| t-test | 24.02 | 752 | < 0.001 |
| TOST Lower | 31.11 | 752 | < 0.001 |
| TOST Upper | 16.93 | 752 | 1 |

Effect Sizes

| | Estimate | SE | C.I. | Conf. Level |
|---|---|---|---|---|
| Raw | 94.7731 | 3.94565 | [88.275, 101.2711] | 0.9 |
| Hedges's g(z) | 0.8745 | 0.04285 | [0.8038, 0.9446] | 0.9 |

Note: SMD confidence intervals are an approximation. See vignette("SMD_calcs").
[1] "==========="
[1] "Troiano60s_W"

Paired t-test

The equivalence test was non-significant, t(752) = 12.93, p = 1
The null hypothesis test was significant, t(752) = 21.319p < 0.01
NHST: reject null significance hypothesis that the effect is equal to zero
TOST: don't reject null equivalence hypothesis

TOST Results

| | t | df | p.value |
|---|---|---|---|
| t-test | 21.32 | 752 | < 0.001 |
| TOST Lower | 29.71 | 752 | < 0.001 |
| TOST Upper | 12.93 | 752 | 1 |

Effect Sizes

| | Estimate | SE | C.I. | Conf. Level |
|---|---|---|---|---|
| Raw | 71.0746 | 3.33392 | [65.584, 76.5651] | 0.9 |
| Hedges's g(z) | 0.7761 | 0.04157 | [0.7076, 0.8442] | 0.9 |

Note: SMD confidence intervals are an approximation. See vignette("SMD_calcs").
